# Disinfection effect of pulsed xenon ultraviolet irradiation on SARS-CoV-2 and implications for environmental risk of COVID-19 transmission

**DOI:** 10.1101/2020.05.06.20093658

**Authors:** Sarah Simmons, Ricardo Carrion, Kendra Alfson, Hilary Staples, Chetan Jinadatha, William Jarvis, Priya Sampathkumar, Roy F. Chemaly, Fareed Khawaja, Mark Povroznik, Stephanie Jackson, Keith S Kaye, Robert M. Rodriguez, Mark Stibich

## Abstract

Prolonged survival of SARS-CoV-2 on environmental surfaces and personal protective equipment (PPE) may lead to these surfaces transmitting disease to others. This article reports the effectiveness of a pulsed xenon ultraviolet (PX-UV) disinfection system in reducing the load of SARS-CoV-2 on hard surfaces and N95 respirators. Chamber slides and N95 respirator material were directly inoculated with SARS-CoV-2 and exposed to different durations of PX-UV disinfection. For hard surfaces, disinfection for 1, 2, and 5 minutes resulted in 3.53 Log_10_, >4.54 Log_10_, and >4.12 Log_10_ reductions in viral load, respectively. For N95 respirators, disinfection for 5 minutes resulted in >4.79 Log_10_ reduction in viral load. We found that PX-UV significantly reduces SARS-CoV-2 on hard surfaces and N95 respirators. With the potential to rapidly disinfectant environmental surfaces and N95 respirators, PX-UV devices are a promising technology for the reduction of environmental and PPE bioburden and to enhance both HCW and patient safety by reducing the risk of exposure to SARS-CoV-2.

## Background

SARS-CoV-2 infected individuals, asymptomatic carriers, and super-shedders readily contaminate the environment which may lead to transmission to other patients and healthcare workers (HCWs) ^1–4^. Shedding can come from both respiratory and fecal secretions ^5–7^. A recent report examining the survival of SARS-CoV-2 in the healthcare environment found that 13/15 (87%) room sites sampled were positive for SARS-CoV-2 by polymerase chain reaction (PCR)^2^. SARS-CoV-2 has been demonstrated to survive on surfaces, such as plastic and steel, for up to 3 days^8^. This extensive spreading and prolonged survival opens the possibility of indirect transmission of SARS-CoV-2 from surfaces, which is consistent with data from prior coronavirus outbreaks such as Severe Acute Respiratory Syndrome (SARS) and Middle Easters Respiratory Syndrome (MERS)^9–12^. Infection clusters of SARS-CoV-2 have been reported, where no direct contact with an infected individual has occurred, but several persons in became infected^13,14^.

These and other data document that the environment poses a risk of SARS-CoV-2 transmission.. Given the fact that is it hard to ensure that manual cleaning/disinfection occurs consistently in healthcare settings^15^ and the fact that cleaning personnel could be at increased risk of exposure to SARS-CoV-2 during their performance of manual cleaning of healthcare facilities, we sought to determine the efficacy of UV-C enhanced environmental disinfection against SARS-CoV-2.

In addition, due to acute shortage of N95 respirators and other personal protective equipment (PPE), healthcare facility personnel have been using a variety of methods (UV-C, hydrogen peroxide, heat, radiation) to disinfect and reuse these PPE^16^. Given that such PPE are repeatedly used by HCWs, it is important to document that such disinfection is effective in reducing SARS-CoV-2 on such PPE, so that HCWs are not exposing themselves to this virus with PPE reuse.

Prevention of healthcare associated infections (HAIs) has been a priority before the COVID-19 pandemic and it is even more important now to prevent SARS-CoV-2 transmission to patients and HCWs. In addition, enhanced SARS-CoV-2 transmission risks exists in other settings such as nursing homes, meat processing plants, prisons and jails, schools, restaurants, and other workplaces.

UV-C has promise as a means of environmental control for SARS-CoV-2. In order to understand the potential of UV-C as a tool in the pandemic, we must first understand the effect of UV-C on SARS-CoV-2 and the necessary operating time to reduce the bioburden of SARS-CoV-2 in the environment. Here we present the results of a laboratory study assessing the efficacy of a full germicidal spectrum UV-C from a pulsed-xenon source (PX-UV) on SARS-CoV-2 on hard surfaces and N95 respirators.

## Methods

### Cells and Virus

Vero E6 cells (VERO C1008, catalog number NR-596, BEI resources) were grown in Dulbecco’s modified essential media (DMEM; Gibco) with 10% heat-inactivated fetal bovine serum (FBS; Gibco) at 37°C with 5% CO_2_.

The SARS-CoV-2 working stock was generated from isolate USA-WA1/2020, obtained from BEI resources (catalog number NR-52281; GenBank accession number: MN985325.1). Virus was passaged one time to generate a master stock, using the following methods. Vero E6 cells were infected at a multiplicity of infection (MOI) of approximately 0.001 in DMEM containing 2% FBS in T150 flasks. Viral supernatant was harvested at 3-days post-infection when the cells exhibited 3+ cytopathic effects, and clarified by low speed centrifugation. This master stock was confirmed to be SARS-CoV-2 via deep sequencing and was stored at <-65°C in 500 ± 50 μL aliquots containing DMEM with 10% FBS. A working stock was generated by infecting Vero E6 cells at a MOI of 0.01 in DMEM containing 2% FBS, in T225 flasks. Viral supernatant was harvested at 3-days post-infection, clarified by low speed centrifugation, and further concentrated by centrifugation at 12,000 × g for 3 hours. Supernatant was removed from the concentrate and the remaining pelleted material pooled to generate the stock used in these experiments. The viral titer was determined to be 1.3 × 10^7^ plaque forming units (PFU)/mL.

### PX-UV Device Testing at Texas Biomedical Research Institute Experimental Design

The procedures and processes utilized to execute the experiment were approved by Texas Biomedical Research Institute institutional review boards. No human participants were involved in this study. The experiments to test the antiviral effects of PX-UV robot model PXUV4D (Xenex Disinfection Services, San Antonio, Texas) on SARS-CoV-2 were performed at the Texas Biomedical Research Institute. Test surfaces (i.e., carriers) included a hard surface (8-well chamber slides) and a soft surface (N95 respirator, 3M Model 1860). Test surfaces were inoculated with 0.020 mL of virus, deposited in a single drop and spread with a pipette tip. Test surfaces were then dried for 55 minutes in a laminar flow hood under ambient conditions. Three carriers per test surface were harvested at time zero to determine the starting viral titer per carrier type, and stored on wet ice while PX-UV exposure occurred for the remaining carriers. There was a 30-minute difference between controls being harvested and all exposures being complete. The robot was placed 1 meter from the test surfaces. (Figure 1) Test surfaces were placed vertically to be parallel with the lamp and exposed in triplicate to the PX-UV device for the specified contact time. Chamber slides were exposed for 1, 2, or 5-minute durations and the N95 respirator carriers were exposed for 5-minute durations. (Table 1) After the exposure period, virus was immediately harvested in 150 μL DMEM supplemented with 2% FBS. Recovered virus was stored on wet ice until processing. Recovered virus was serially diluted (100 μL was used to prepare a 1:1 dilution and 50 μL was used to prepare serial ten-fold dilutions). This material was subjected to plaque assay as described below. Viral titers were determined as PFU/mL in starting material harvested from the carriers. Data was analyzed by authors RC, KA, and HS.

**Figure 1.**
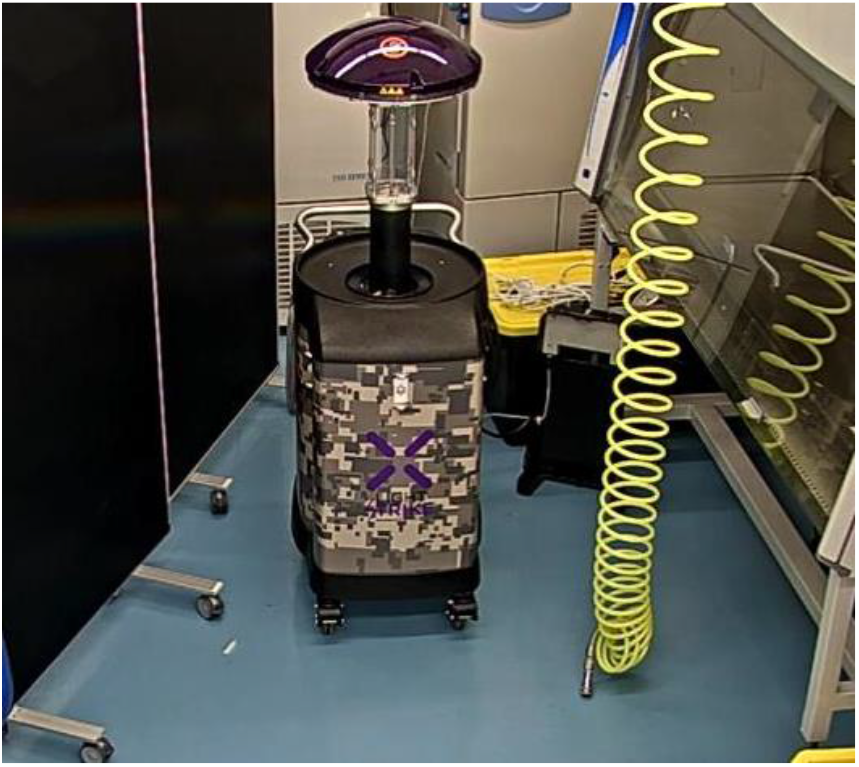
PX-UV Device

**Table 1.** Experimental Design

| Test Microorganism | Test Device | Treatment Distance | Test Surface | Contact Time | Number |
| --- | --- | --- | --- | --- | --- |
| SARS-CoV-2 | PX-UV Device | 1 meter | Chamber Slide | 0 minutes | 3 |
|  |  |  | Chamber Slide | 1 minute | 3 |
|  |  |  | Chamber Slide | 2 minutes | 3 |
|  |  |  | Chamber Slide | 5 minutes | 3 |
|  |  |  | N95 Respirator | 0 minutes | 3 |
|  |  |  | N95 Respirator | 5 minutes | 3 |

\*SARS-CoV2: Severe Acute Respiratory Syndrome Coronavirus 2, PX-UV: Pulsed Xenon Ultraviolet

### Determination of Viral Titers

Viral titers were determined by plaque assay using a methylcellulose and crystal violet assay^17,18^. Vero E6 cells were seeded in 12-well tissue culture plates in DMEM (Gibco) with 10% heat-inactivated FBS (Gibco) at a density of 2 × 10^5^ cells per well. Positive control samples (i.e., material from slide and N95 respirator with no UV exposure) were serially diluted ten-fold and test samples were diluted 1:1 and serially ten-fold. Dilutions were prepared in DMEM containing 2% FBS. Media were removed from the plates and 100μL of each dilution was added to the corresponding well in duplicate. A negative control plate was prepared as well. The plates were incubated for one hour at 37°C with 5% CO_2_, with constant rocking. After incubation, media were removed from the wells and a 2mL primary overlay consisting of DMEM with 2% heat-inactivated FBS and 30% methylcellulose (Sigma-Aldrich) was added. Plates were then incubated at 37°C with 5% CO_2_ for three days. After three days, the overlay was removed and 10% neutral buffered formalin (Sigma-Aldrich) was added to each well to fix the cells. After fixation, formalin was removed and the plates were washed in 1X PBS (Gibco). To stain the plates, approximately 500uL of crystal violet stain (Ricca Chemical Company) were added to each well and plates were incubated at room temperature for 10 minutes. Then, the plates were washed in fresh water and allowed to air dry before plaques were counted to determine a final titer. (Figure 2)

**Figure 2.**
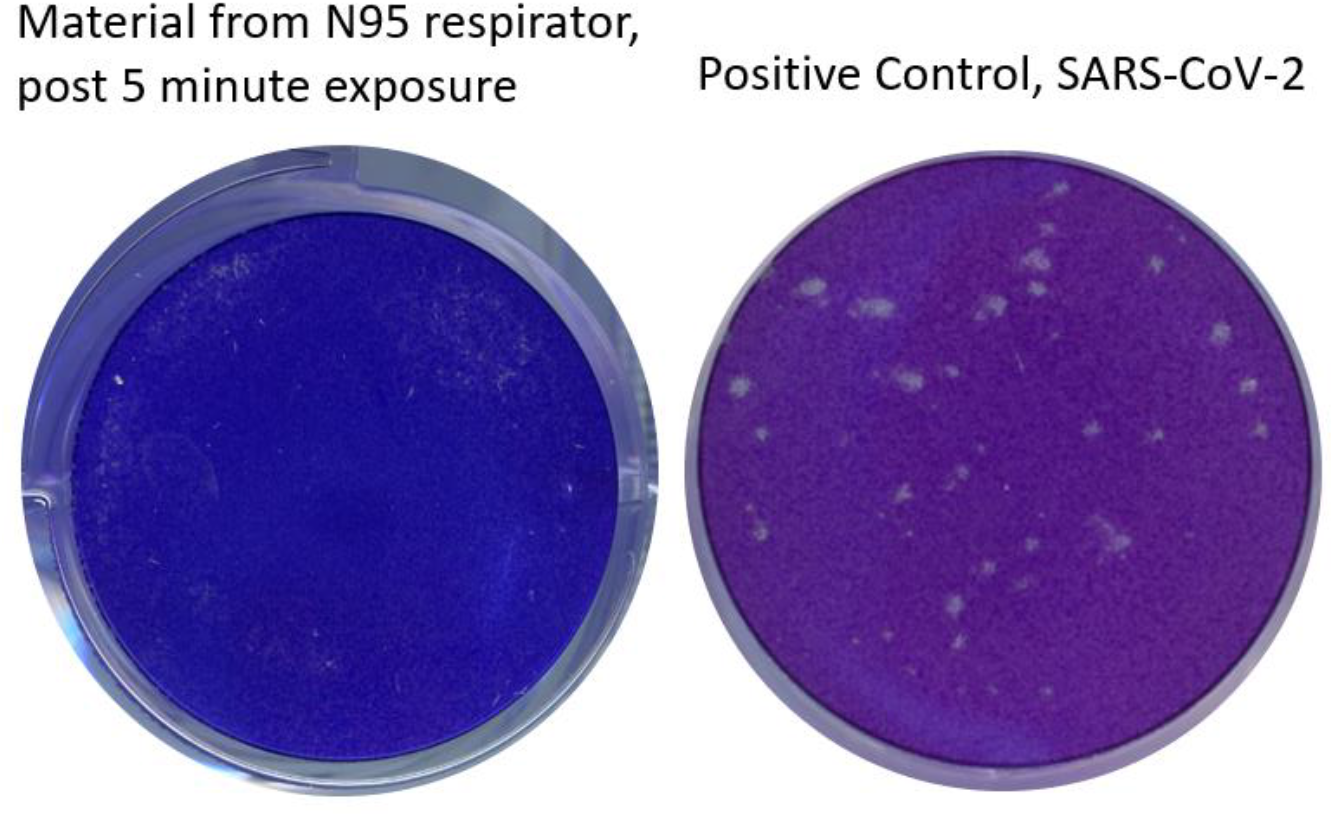
Images of Stained Cells Inoculated with Materials from Exposed and Control Carrier, from N95 Respirator Testing

## Results

The controls for the hard surfaces carriers averaged a titer of 6.20 PFU/mL (Log_10_). In contrast, the PX-UV exposed hard surfaces had 2.67 PFU/mL (Log_10_) (3.56 Log_10_ reduction,99.97%), <1.66 PFU/mL (Log_10_)(>4.54 Log_10_ reduction, 99.997%), and <2.08 PFU/mL (Log_10_)(>4.12 Log_10_ reduction, 99.992%) at 1, 2 and 5-minute PX-UV exposure, respectively (Table 2). The detection threshold of the experimental methods was 1.3 PFU/mL (Log_10_) and that value was inserted when the levels of SARS-CoV-2 on the carriers were undetectable.

**Table 2.** Impact of PX-UV on SARS-CoV-2 Inoculated onto Hard Surfaces

| Test Surface | UV-C Exposure Time [minutes] | Viral Titer [PFU/mL (Log<sub>10</sub>)]<br>per Carrier |  |  | Average | % reduction | Log<sub>10</sub> Reduction Relative to Respective Controls |
| --- | --- | --- | --- | --- | --- | --- | --- |
|  |  | 1 | 2 | 3 |  |  |  |
| Slide | 0 | 6.04 | 6.28 | 6.28 | 6.20 | N/A | N/A |
| Slide | 1 | 3 | 2.45 | 2.56 | 2.67 | 99.97% | 3.53 |
| Slide | 2 | 2.38 | <1.3 | <1.3 | <1.66 | >99.997% | >4.54 |
| Slide | 5 | <1.3 | 2.15 | 2.8 | <2.08 | >99.992% | >4.12 |

\*UV-C: Ultraviolet C light, PFU: Plaque forming units

Next, we evaluated the impact of PX-UV on SARS-CoV-2 inoculated on N95 respirators. Control titers averaged at 6.35 PFU/mL (Log_10_). Inoculated N95 respirators exposed to 5 minutes of PX-UV showed <1.56 PFU/mL (Log_10_), or a >4.79 Log_10_ reduction (99.998%)(Table 3). The detection threshold of the experimental methods was 1.3 PFU/mL (Log_10_) and that value was inserted when the levels of SARS-CoV-2 on the carriers were undetectable.

**Table 3.** Impact of PX-UV on SARS-CoV-2 Inoculated on N95 Respirators

| Test Surface | UV-C Exposure Time [minutes] | Viral Titer [PFU/mL (Log<sub>10</sub>)]<br>per Carrier |  |  | Average | % reduction | Log<sub>10</sub> Reduction Relative to Respective Controls |
| --- | --- | --- | --- | --- | --- | --- | --- |
|  |  | 1 | 2 | 3 |  |  |  |
| N95 Respirator | 0 | 6.84 | 6.04 | 6.18 | 6.35 | N/A | N/A |
| N95 Respirator | 5 | <1.3 | <1.3 | 2.08 | <1.56 | >99.998% | >4.79 |

\*UV-C: Ultraviolet C light, PFU: Plaque forming units

## Discussion

The results from our study demonstrate that the rapid disinfection times from PX-UV devices can effectively reduce the viable load of SARS-CoV-2 in a laboratory setting on both chamber slides and N95 respirators. To our knowledge, this PX-UV device is the first no-touch disinfection system to show efficacy directly against SARS-CoV-2 on hard surfaces. UV-C has been the most common method of PPE disinfection in response to the pandemic, despite conflicting data regarding its efficacy. Our study is also the first to demonstrate that PX-UV specifically is effective in reducing SARS-CoV-2 on N95 respirators. The results of tests demonstrating that disinfection with PX-UV will not impact the fit or function of the respirators are available from the respirator manufacturer^19^.

Use of PX-UV is not a novel concept and has been deployed for HAI prevention, including multidrug-resistant organisms (MDROs) such as methicillin-resistant *Staphylococcus aureus* (MRSA) and *Clostridioides difficile* ^20–23^. The device is most commonly used during the terminal cleaning of patient rooms, after manual disinfection using an EPA-registered disinfectant. With SARS-CoV-2, there will be additional target areas for disinfection. Given that emergency departments (EDs) and SARS-CoV-2 testing centers are the primary sites for triage and evaluation of suspected SARS-CoV-2 patients, use of PX-UV should be considered throughout these areas; including triage, patient rooms, radiology suites, and patient bathrooms. Considering the potential for secondary transmission, non-patient care areas (i.e., lobbies, waiting rooms, staff break rooms, cafeterias and staff on-call rooms) should be considered for disinfection as well. Portable medical equipment (PME) also should also be considered as possible vectors for transmitting SARS-CoV-2. Equipment such as mobile workstations, vital signs machines, wheel chairs, and intravenous (IV) pumps, can become heavily contaminated with routine use^24,25^. Disinfection of PME with a PX-UV device resulted in a 94% reduction in bacterial load^26^. Our demonstration that brief cycles of PX-UV disinfection are effective in decreasing SARS-CoV-2 attests the feasibility of its use in these settings.

Our study has several limitations. We did not evaluate the direct effect of PX-UV on existing healthcare environmental contamination, but rather high virion concentration in a laboratory setting. Our inoculum exceeds the level of SARS-CoV-2 contamination that would be seen in a routine clinical healthcare environment. It is likely that in such clinical environments, the impact of the PX-UV in reducing environmental bioburden would be even greater.

The results from our study cannot be generalized to other UV light sources because UV-C from a PX-UV system is fundamentally different from that produced by other UV disinfection systems that rely on low-pressure mercury vapor lamps or light emitting diode (LED) sources^27^. UV-C from a PX-UV system produces broad-spectrum wavelength light that covers the entire germicidal UV spectrum, from 200-280 nanometers (nm)^28^, potentially creating more viricidal effect than the wavelengths produced by these other narrow spectrum sources^29^.

## Conclusion

We found that PX-UV significantly reduces SARS-CoV-2 on hard surfaces and N95 respirators. With the potential to rapidly disinfectant environmental surfaces and N95 respirators, PX-UV devices are a promising technology for the reduction of environmental and PPE bioburden and to enhance both HCW and patient safety by reducing the risk of exposure to SARS-CoV-2.

## Data Availability

All data generated or analysed during this study are included in this published article

## References

1. Santarpia JL, Rivera DN, Herrera V, et al. Transmission Potential of SARS-CoV-2 in Viral Shedding Observed at the University of Nebraska Medical Center. medRxiv 2020:2020.03.23.20039446.

2. Ong SWX, Tan YK, Chia PY, et al. Air, Surface Environmental, and Personal Protective Equipment Contamination by Severe Acute Respiratory Syndrome Coronavirus 2 (SARS-CoV-2) From a Symptomatic Patient. Jama 2020;323:1610–2.

3. Guo ZD, Wang ZY, Zhang SF, et al. Aerosol and Surface Distribution of Severe Acute Respiratory Syndrome Coronavirus 2 in Hospital Wards, Wuhan, China, 2020. Emerging infectious diseases 2020;26.

4. Gudbjartsson DF, Helgason A, Jonsson H, et al. Spread of SARS-CoV-2 in the Icelandic Population. The New England journal of medicine 2020.

5. Gu J, Han B, Wang J. COVID-19: Gastrointestinal Manifestations and Potential Fecal–Oral Transmission. Gastroenterology 2020;158:1518–9.

6. Jiang X, Luo M, Zou Z, Wang X, Chen C, Qiu J. Asymptomatic SARS-CoV-2 infected case with viral detection positive in stool but negative in nasopharyngeal samples lasts for 42 days. Journal of Medical Virology 2020;n/a.

7. Xiao F, Tang M, Zheng X, Liu Y, Li X, Shan H. Evidence for Gastrointestinal Infection of SARS-CoV-2. Gastroenterology 2020;158:1831–3.e3.

8. van Doremalen N, Bushmaker T, Morris DH, et al. Aerosol and Surface Stability of SARS-CoV-2 as Compared with SARS-CoV-1. The New England journal of medicine 2020.

9. Dowell SF, Simmerman JM, Erdman DD, et al. Severe Acute Respiratory Syndrome Coronavirus on Hospital Surfaces. Clinical Infectious Diseases 2004;39:652–7.

10. Khan RM, Al-Dorzi HM, Al Johani S, et al. Middle East respiratory syndrome coronavirus on inanimate surfaces: A risk for health care transmission. American journal of infection control 2016;44:1387–9.

11. Otter JA, Donskey C, Yezli S, Douthwaite S, Goldenberg SD, Weber DJ. Transmission of SARS and MERS coronaviruses and influenza virus in healthcare settings: the possible role of dry surface contamination. The Journal of hospital infection 2016;92:235–50.

12. Xiao S, Li Y, Wong T-W, Hui DSC. Role of fomites in SARS transmission during the largest hospital outbreak in Hong Kong. PloS one 2017;12:e0181558-e.

13. Cai J, Sun W, Huang J, Gamber M, Wu J, He G. Indirect Virus Transmission in Cluster of COVID-19 Cases, Wenzhou, China, 2020. Emerging infectious diseases 2020;26.

14. Moriarty LF, Plucinski MM, Marston BJ, et al. Public Health Responses to COVID-19 Outbreaks on Cruise Ships - Worldwide, February-March 2020. MMWR Morbidity and mortality weekly report 2020;69:347–52.

15. Carling PC, Parry MM, Rupp ME, Po JL, Dick B, Von Beheren S. Improving cleaning of the environment surrounding patients in 36 acute care hospitals. Infection control and hospital epidemiology: the official journal of the Society of Hospital Epidemiologists of America 2008;29:1035–41.

16. Strategies for Optimizing the Supply of N95 Respirators: Contingency Capacity Strategies. 2020. (Accessed March 23, 2020, at https://www.cdc.gov/coronavirus/2019-ncov/hcp/respirators-strategy/contingency-capacity-strategies.html.)

17. Wang S, Sakhatskyy P, Chou T-HW, Lu S. Assays for the assessment of neutralizing antibody activities against Severe Acute Respiratory Syndrome (SARS) associated coronavirus (SCV). Journal of Immunological Methods 2005;301:21–30.

18. Tan ELC, Ooi EE, Lin C-Y, et al. Inhibition of SARS coronavirus infection in vitro with clinically approved antiviral drugs. Emerging infectious diseases 2004;10:581–6.

19. Decontamination Methods for 3M N95 Respirators. 2020. (Accessed April 16, 2020, at https://multimedia.3m.com/mws/media/1824869O/decontamination-methods-for-3m-n95-respirators-technical-bulletin.pdf.)

20. Sampathkumar P, Folkert C, Barth JE, et al. A trial of pulsed xenon ultraviolet disinfection to reduce Clostridioides difficile infection. American journal of infection control 2019;47:406–8.

21. Levin J, Riley LS, Parrish C, English D, Ahn S. The effect of portable pulsed xenon ultraviolet light after terminal cleaning on hospital-associated Clostridium difficile infection in a community hospital. American journal of infection control 2013;41:746–8.

22. Haas JP, Menz J, Dusza S, Montecalvo MA. Implementation and impact of ultraviolet environmental disinfection in an acute care setting. American journal of infection control 2014;42:586–90.

23. Simmons S, Morgan M, Hopkins T, Helsabeck K, Stachowiak J, Stibich M. Impact of a multihospital intervention utilising screening, hand hygiene education and pulsed xenon ultraviolet (PX-UV) on the rate of hospital associated meticillin resistant Staphylococcus aureus infection. Journal of Infection Prevention 2013.

24. Jinadatha C, Villamaria FC, Coppin JD, et al. Interaction of healthcare worker hands and portable medical equipment: a sequence analysis to show potential transmission opportunities. BMC infectious diseases 2017;17:800.

25. John A, Alhmidi H, Cadnum JL, Jencson AL, Donskey CJ. Contaminated Portable Equipment Is a Potential Vector for Dissemination of Pathogens in the Intensive Care Unit. Infect Control Hosp Epidemiol 2017;38:1247–9.

26. Reid D, Ternes K, Winowiecki L, et al. Germicidal irradiation of portable medical equipment: Mitigating microbes and improving the margin of safety using a novel, point of care, germicidal disinfection pod. American journal of infection control 2020;48:103–5.

27. Kowalski W. Ultraviolet Germicidal Irradiation Handbook: UVGI for Air and Surface Disinfection. Berlin, Heidelberg: Springer Berlin Heidelberg; 2009.

28. Schaefer R, Grapperhaus M, Schaefer I, Linden K. Pulsed UV lamp performance and comparison with UV mercury lamps. Journal of Environmental Engineering and Science 2007;6:303–10.

29. Beck SE, Wright HB, Hargy TM, Larason TC, Linden KG. Action spectra for validation of pathogen disinfection in medium-pressure ultraviolet (UV) systems. Water Res 2015;70:27–37.

